# pyPOCQuant - A tool to automatically quantify Point-Of-Care Tests from images

**DOI:** 10.1101/2020.11.08.20227470

**Authors:** Andreas P. Cuny, Fabian Rudolf, Aaron Ponti

## Abstract

Lateral flow Point-Of-Care Tests (POCTs) are a valuable tool for rapidly detecting pathogens and the associated immune response in humans and animals. In the context of the SARS-CoV-2 pandemic, they offer rapid on-site diagnostics and can relieve centralized laboratory testing sites, thus freeing resources that can be focused on especially vulnerable groups. However, visual interpretation of the POCT test lines is subjective, error prone and only qualitative. Here we present pyPOCQuant, an open-source tool implemented in Python 3 that can robustly and reproducibly analyze POCTs from digital images and return an unbiased and quantitative measurement of the POCT test lines.

## 1. Motivation and significance

Point of care tests (POCT) and rapid diagnostic tests (RTD) are a cost-effective and instrument-free alternative to molecular laboratory testing used for a wide variety of diseases and the associated immune response, especially in times or regions where resources are limited [1]. Most POCTs are Immunochromatographic Lateral Flow Assays (LFA) or similar, that use one binding protein coated on a membrane and a second protein fused to a colloidal Gold entity to report a positive reaction as a visible color line [2]. The main components of an LFA (Fig. 1a) are, therefore, a nitrocellulose membrane with test and control lines, a sample pad where a conjugate is stored, and the absorption pad [3].

**Figure 1:**
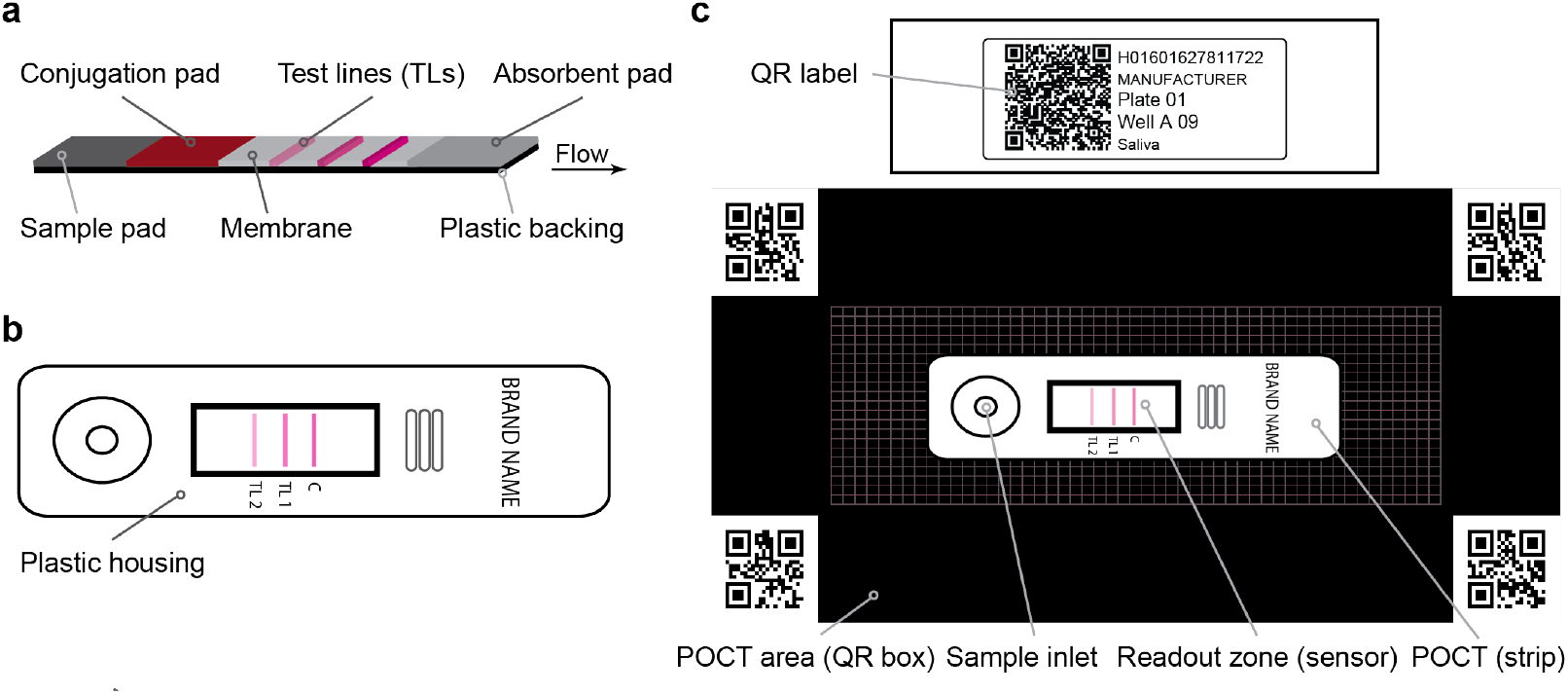
Typical design of lateral flow based POCTs. a) Configuration of LFA. b) POCT schematic enclosing the LFA. c) Example QR code encoding patient metadata (details in text) with POCT area and POCT positioning on template (Suppl. Fig. S1).

POCTs are commonly used for Streptococcus diagnostics and have been used to combat the epidemics of HIV, TB, hepatitis, dengue fever, and malaria. They all use binding proteins specific for the pathogen as their test line and color conjugate. Analogously, antibody assays reveal whether a patient’s immune system has already developed antibodies (i.e., IgM, IgA, or IgG) against past viral infections. A sample is taken using a swab (e.g., in Streptococcus diagnostic), a small amount of blood (e.g., in antibody detection) or similar, and mixed with running buffer either outside or directly on the sample pad of the POCT. In the best case, POCTs deliver fast readouts (typically within 15 minutes) close to the patient and allow for fast reactions with a potential advantage for the patient and the whole local community [4].

To be of use in the global SARS-CoV-2 pandemic, POCTs must fulfill the standards set by the WHO (≥ 80% sensitivity and ≥ 97 − 99.5% specificity) [5, 6]. These specifications hold both for the diagnostic of an ongoing viral infection and the detection of past infections. A plethora of manufacturers provide tests that they claim to fulfill these specifications, but we argue that they will require comprehensive characterization and comparison of their analytical and clinical performance [7, 8].

LFA and similar devices are scored visually and yield a dichotomous (positive/negative) readout. Experimentally, the analytical limit of detection can be determined using a titration of (repeated) measurements of purified proteins (such as monoclonal antibodies as a surrogate for blood IgG detection) or in proteins from the pathogen of interest that are expressed in cell lines and purified (i.e. Spike protein of SARS-CoV-2) [9]. However, they do have the potential for a quantitative readout using dedicated readers, lab scanner, or image-based analysis [10, 11, 12, 13]. While there have been efforts to use smartphones as readers in combination with 3D-printed adapters and holders [14, 15, 16, 17, 18, 19, 20], no freely available pipeline is available to analyze POCTs from large numbers of images in an automated way.

Here, we present pyPOCQuant, an open-source tool for the large-scale, quantitative and automated analysis of large numbers of POCTs from a variety of different manufacturers independent of design. pyPOCQuant processes images of POCTs taken on digital cameras and returns both a binary result (positive vs. negative) for the test and control lines, and a quantitative readout of each test line. These robust and reproducible readouts are well suited for studying the temporal evolution of the patient’s immune response to the infectious agent, or as a quick diagnostics tool for detecting a pathogen. As such, pyPOCQuant helps with the validation and characterisation of different POCTs, but it can also make the use of these assays more reliable.

## 2. Software description

pyPOCQuant is a cross platform tool written in Python 3 and implemented as a stand-alone computer vision library. It is available under the open-source license GPLv3 on the Python Package Index (`pip`) and on GitHub, both as source or as compiled stand-alone executable. Extensive documentation (including example scripts and data) can be found on readthedocs.org and bundled with the application. A quick start guide highlights the steps required to properly set up image acquisition and to run a successful analysis. The user manual describes its usage in full detail.

### 2.1. Software Architecture

Depending on the workflow, needs and preferences of the user, pyPOCQuant can be launched from the command line interface (CLI) using the `pyPOCQuant.py` main script, or run as a module from a Jupyter notebook. We also developed pyPOCQuantUI, a user-friendly graphical user interface (GUI) and desktop application that leverages pyPOCQuant while massively facilitating the definition of essential parameters needed for the analysis. pyPOCQuantUI is implemented in Python 3 using the PyQt5 library and can be run from source or as a stand-alone, executable desktop application. Figure 2 displays a flowchart of the operations performed by pyPOCQuant. In short, for every image in a folder, pyPOCQuant extracts patient and metadata information, segments and processes the POCT, extracts its sensor area, locates and quantifies the test-line signals. All results are stored in a data frame that can be used for further statistical analysis. The next sections will discuss these operations in detail.

**Figure 2:**
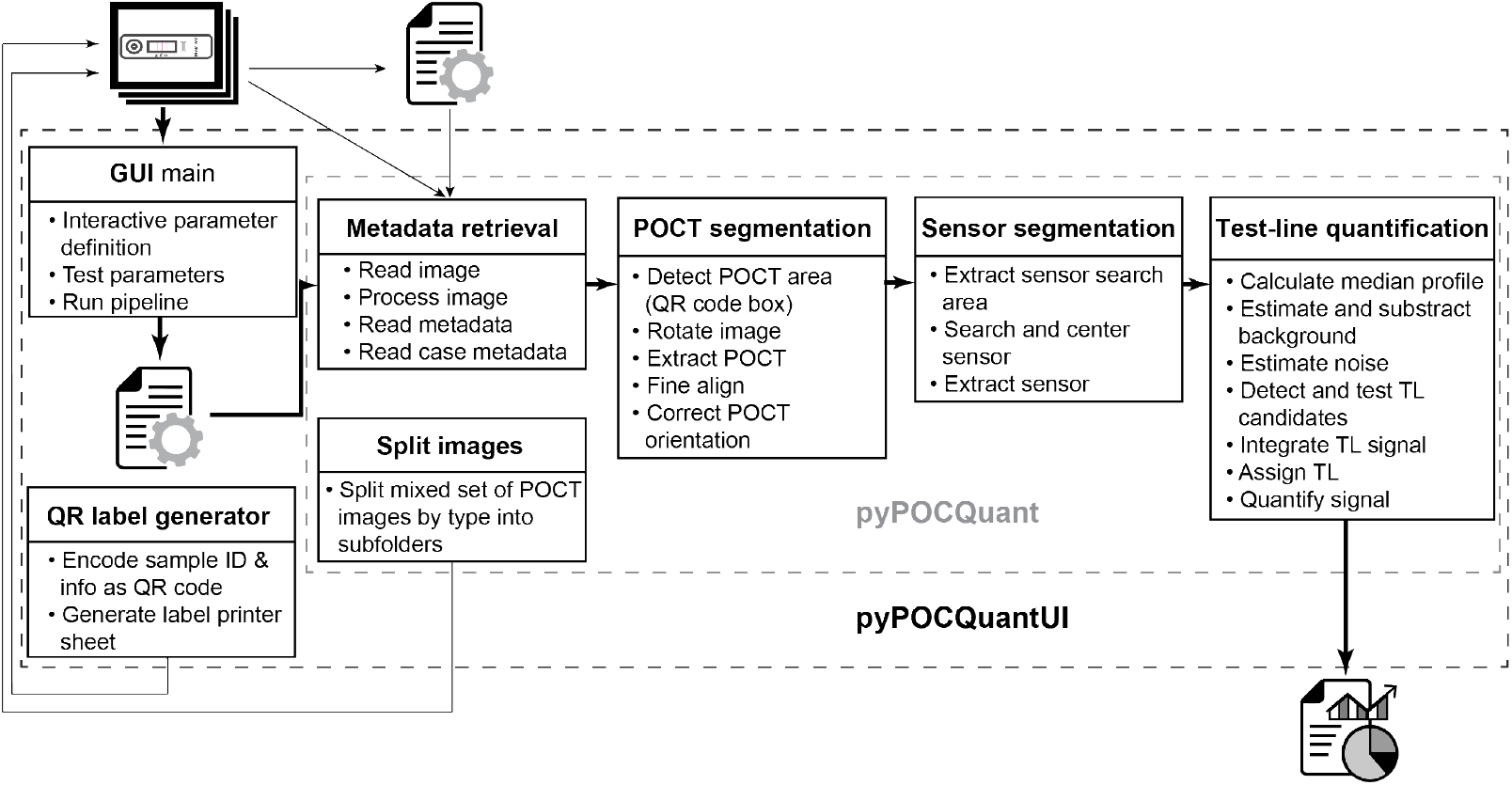
pyPOCQuant architecture overview. Bold arrows depict the main flow trough the tool using the GUI. Regular arrows depict the CLI usage and optional ways of interaction.

### 2.2. Software Functionalities

The core of pyPOCQuant is a pipeline of operations that can be configured and started either from the CLI or GUI. Parameters are passed to the pipeline via a configuration file.

#### 2.2.1. Pipeline

The pipeline processes all images in a user-defined input folder in parallel, and the number of parallel tasks can be defined at run time (Figure 2). Results are compiled in a common data frame and saved as a comma-separated-value file in a user-defined output folder. The following sections describe the operations that are performed on each image independently.

##### Metadata retrieval

Current image from a series of formats (JPEG and various RAW image formats, specifically Nikon NEF, Sony ARW and Canon CR2) is read from disk. Image metadata such as acquisition date and time, exposure time, focal length, ISO, are read and stored in the data frame. Use of the provided template (Suppl. Fig. S1) for image acquisition ensures that patient information stored in a QR code placed on the template can be automatically retrieved and associated directly to the image.

##### POCT segmentation

Additional QR codes encoding spatial information make the process of segmentation more robust. These QR codes ensure that the image was acquired in the expected orientation. If this is not the case, a coarse initial rotation (in steps of 90 degrees) is performed. The positional QR codes delimit the area where the POCT is placed on the template (Suppl. Fig. S1). The POCT area is large enough to guarantee that POCTs of different shapes and sizes can be contained and robustly identified. The POCT area is extracted and finely rotated to be parallel to the image axes. The POCT itself is segmented and rotated to be perfectly aligned to the image axes. Since it may happen that the POCT was incorrectly rotated by 180 degrees when placed on the template, optional feature detection and OCR are used to ensure that the inlet and the control TL are on the expected side of the image.

##### Readout zone (sensor) segmentation

Once the properly aligned POCT area has been segmented from the image, a candidate sensor window of size `sensor_search_area` and centered at the `sensor_center` coordinates is extracted. Using the control TL as a visual anchor and its expected position within the sensor area as defined in `peak_expected_relative_location`, the search area is properly center around the `sensor_center` coordinates and the sensor of given `sensor_size` is cropped for further analysis.

##### Test lines (TL) quantification

As the first step in the detection and analysis of the TL signals, a one-dimensional signal profile is calculated as the median intensity along the short side of the sensor window. This increases the robustness of the analysis by reducing variations in the TL signals due to inhomogeneous reactions of the bands (Figure 3). Also, since the edges of the readout zone often show some impurities, `sensor_border` rows and columns of the sensor image are ignored when calculating the signal profile.

**Figure 3:**
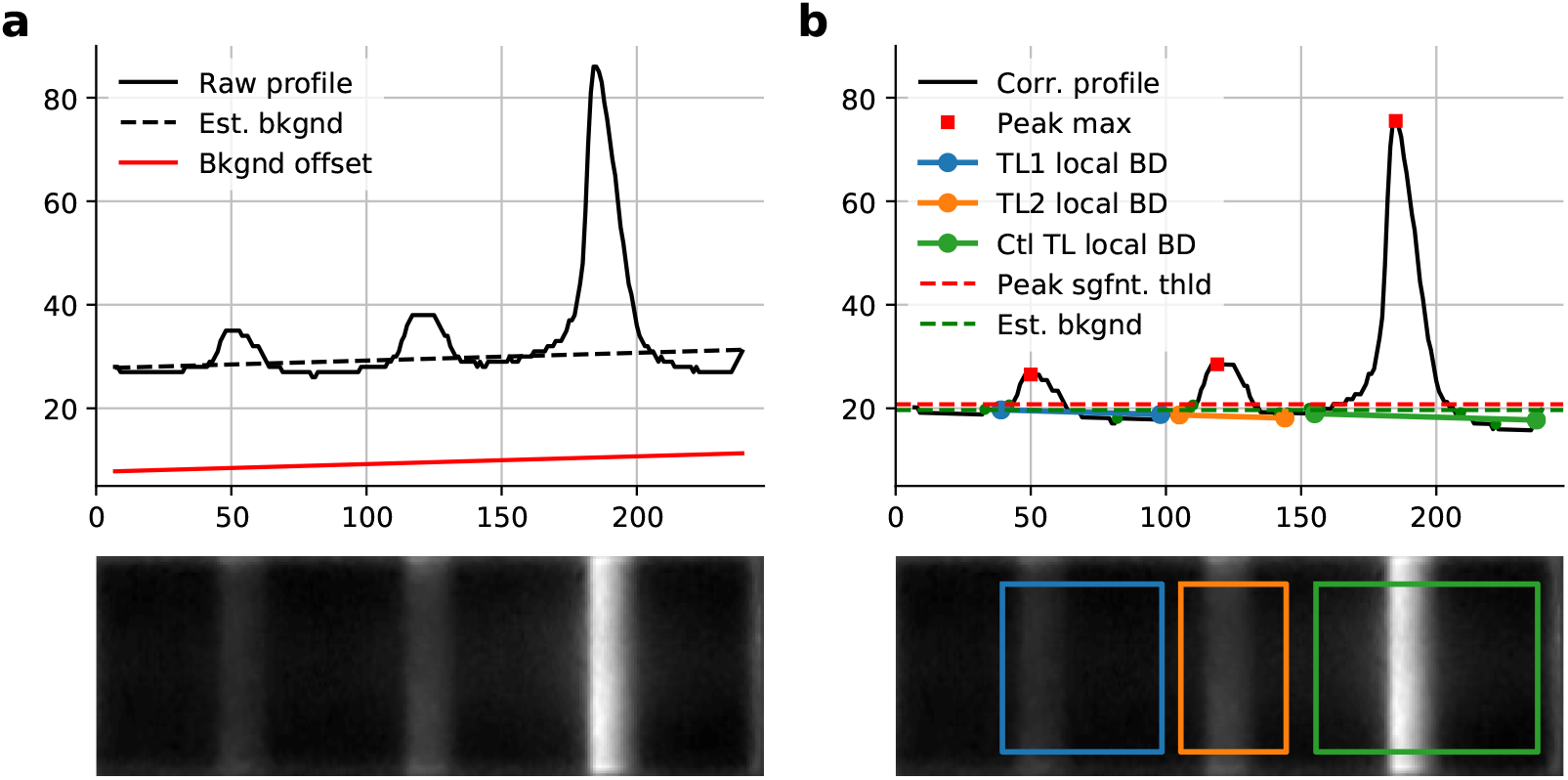
Test line quantification. a) Background estimation (Est. bkgnd) and fixed offset (Bkgnd offset) from raw median intensity profile. b) Background corrected intensity profile (Corr. profile) with peak maxima (red squares) and TL bounds (BD: blue, orange, green circle and lines) as well as peak significance threshold (red dashed line) and the estimated background (black dashed line). The raw grey-scale image plotted below. The colored squares depict all the pixels considered for the signal quantification.

A robust linear regressor (Huber regressor, [21]) is used to fit the background level, that often shows an increasing or decreasing trend along the profile. Local minima in the profile are iteratively used to build a background line that is subtracted from the profile. A small constant subtracted from the background line guarantees that no values of the subtracted profile are negative. Next, the profile is low-pass filtered with a small Gaussian kernel (*σ* = 1) to clean spurious signal variations, and a local max detector tuned on the size of the expected TLs provides candidate location for the test lines. To avoid scoring local maxima due to noisy signal variations, a thresholding step is performed to make sure that only significant peaks are considered for further analysis. This threshold is calculated from the median variation of the background local minima and the multiplicative factor `sensor_thresh_factor`. The remaining local maxima are scored by their relative proximity to the `peak_expected_relative_location` and assigned to one of the `T1`, `T2` or `Ctl` TLs. Last, each TL signal is calculated as the integral of the profile from background level to background level on the two sides of the peak. This decision was taken after observing that a binary segmentation of the TLs could result in a significant variation in the extent of the TLs across images, and thus increase the error rate in the detection of absolute and relative signal strengths.

Once all TLs have been extracted and their absolute signals calculated, the binary readout is recorded. A TL is present if a significant peak could be extracted and its relative signal strength is normalized against the absolute signal of the control TL. Absolute values are also reported in the results data frame to make sure that intra-batch variability can be accounted and corrected for. If a control TL cannot be found and extracted from the sensor area, the test is recorded but is labeled as invalid.

##### Outputs

The results for each image are collected into a data frame and saved into the output folder. The structure of the results file is described in the user manual. Result images and a full log of the process are saved along. The verbosity of the log and the creation of additional quality control images can be controlled in the settings.

#### 2.2.2. Patient sample QR code label generator

When working with many POCTs from many patients and/or replicates, it is essential to link samples and patients unequivocally. Typically, an identification number is associated with each sample. To facilitate the association of patient and test metadata with the corresponding POCT analysis readouts, we have added a QR-code generation tool to pyPOCQuantUI that allows flexible printing of labels in the format `SAMPLEID-MANUFACTURER-PLATE-WELL-USERDATA`. The label can contain additional information such as sample id, manufacturer names, identification for sample replicates, organisation in standardized plates (especially 96-well plates) and a short free text. With the default page and label settings the full label should not exceed 60 characters. The generated labels are compiled in one PDF file and can be printed with ordinary printers and placed on the template next to the POCT (Fig. 1c). pyPOCQuant will read this information and add it to the corresponding analysis results in the data frame.

## 3. Illustrative Examples

### 3.1. Example usage with the GUI

pyPOCQuantUI (Figure 4) facilitates the definition of the analysis parameters by offering interactive tools and visual cues. Also, its immediate feedback allows checking results and fine-tuning parameters quickly. The user manual describes all functionalities of the interface in full detail.

**Figure 4:**
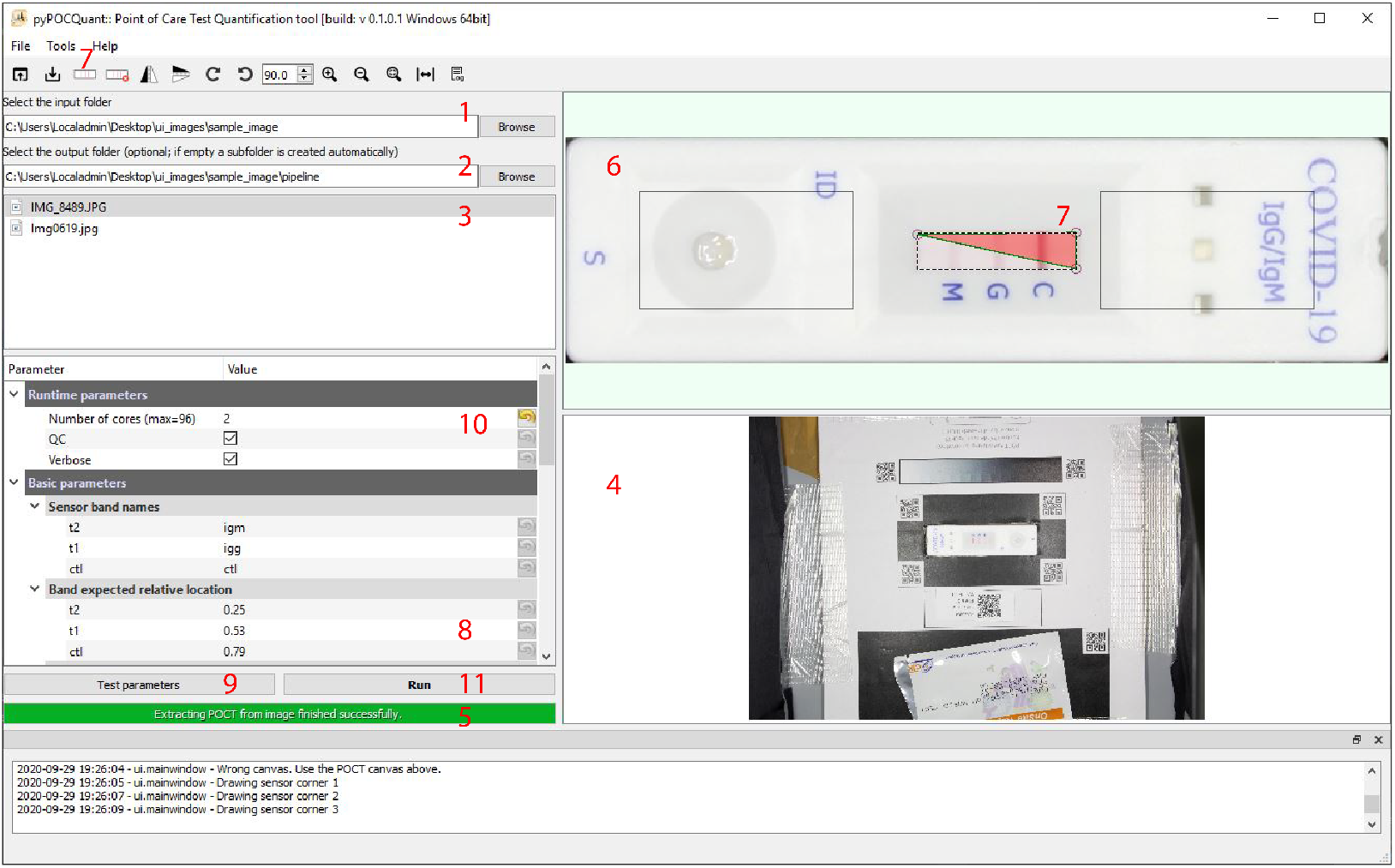
pyPOCQuantUI (GUI): interactively defining basic analysis parameters. The numbers correspond to the text.

#### Configuration and working folders

pyPOCQuantUI starts with a default configuration already loaded. Alternatively, configuration files can be loaded from the file menu, the toolbar, or simply clicking on a `*.conf` file in the input file list. A new project is started by selecting an image folder (1). By default, the target results folder will target a `pipeline` subfolder in the input folder, but it can be changed (2). The files contained in the input folder are listed in (3).

#### Definition of basic parameters

Selecting one of the images in the list will display it on the right (4) and trigger the POCT extraction. Its progress is shown in (5). The extracted POCT is displayed in (6). By clicking on the `draw_sensor_icon` in the toolbar, the user can draw a polygon that connects the four corners of the readout zone of the POCT (7). This interactive sensor can be edited and deleted. Mouse wheel zooming or using the zoom icons facilitate the accurate drawing of the interactive sensor. The sensor polygon also features three vertical test lines that need to be aligned with the test lines to defined their expected relative position. The vertical lines’ position can be changed with the mouse or by editing their values in the parameter tree (8).

#### Test and run pipeline

At this stage, all the basic parameters are set and can be tested by hitting the `Test parameters` button (9). It is recommended (and activated by default) to enable creation of quality control images and verbose logging during testing. When the parameters deliver satisfying results, one can increase the number of parallel workers (10) and start batch-processing all images in the input folder in parallel (11). The `quantification_data.csv` result file, log files, and, optionally, the quality control images will be stored in the output folder.

#### Advanced topics

pyPOCQuant’s advanced parameters and detailed discussion of the results can be found in the User manual or in the Quick-start guide, which also summarizes a few common issues and suggests ways to resolve them. Worth mentioning is the `Issue` column in the result data frame that contains a non-zero error code if the analysis of the corresponding image could not be completed successfully. The issue codes can be found in the manual.

### 3.2. Example usage with the CLI

Running pyPOCQuant from the CLI is best suited when automating the processing of large amounts of images and folders. A configuration with default values is created (Listing 1) and the parameters are modified appropriately. The easiest approach is to use pyPOCQuantUI for this purpose, but it could also be done with other tools, such as Fiji (as described in the manual). To create a default configuration from the CLI, use the `‘-c’` flag of pyPOCQuant.py.

Listing 1: Create a new default settings file

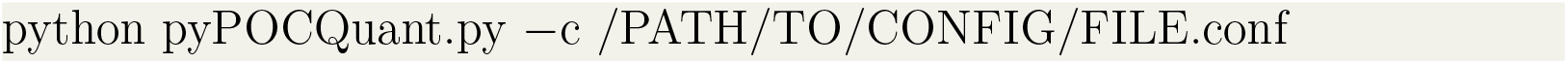

Once the configuration file is ready, a full study can be started by running pyPOCQuant on a full folder of images (Listing 2). The analysis is performed in parallel, and the number of concurrent tasks can be defined by the `‘-w’` (`--workers`) argument. The maximum number of workers allowed corresponds to the maximum number of CPU cores.

Listing 2: Usage of pyPOCQuant with the CLI on an image folder with given settings file and number of workers

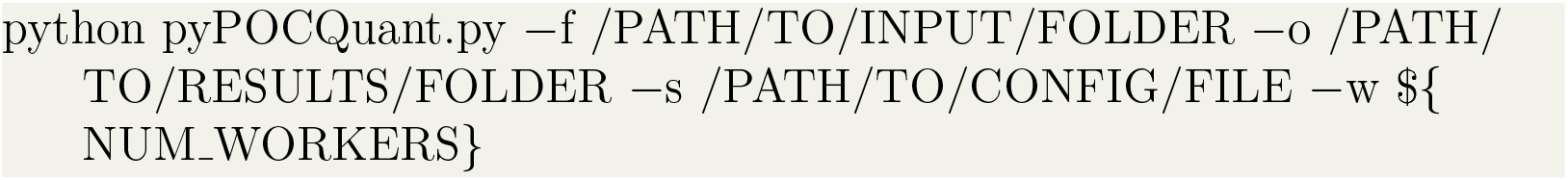

The argument `‘-f’` is the path to the folder that contains all images for a given camera and manufacturer to be processed; `‘-o’` is the path where the `‘quantification_data.csv’` result file, log and quality control images are saved (optional, if omitted the path defaults to a `pipeline` sub-folder in the input folder); `‘-s’` sets the path to the configuration file to be used for this analysis; `‘-w’` is the number of parallel processes; e.g. `8`.

### 3.3. Example usage with Jupyter notebook

Finally, pyPOCQuant can be run from a Jupyter notebook. A full demonstration notebook along with example images can be found in the repository and the online documentation. Documentation of the parameters and outputs can be found in the User manual. Importing of `run_pipeline` and `default_settings` along with the path to images are enough to run pyPOC-Quant from a Jupyter cell (Listing 3).

Listing 3: Minimal example of usage of pyPOCQuant in a Jupyter notebook

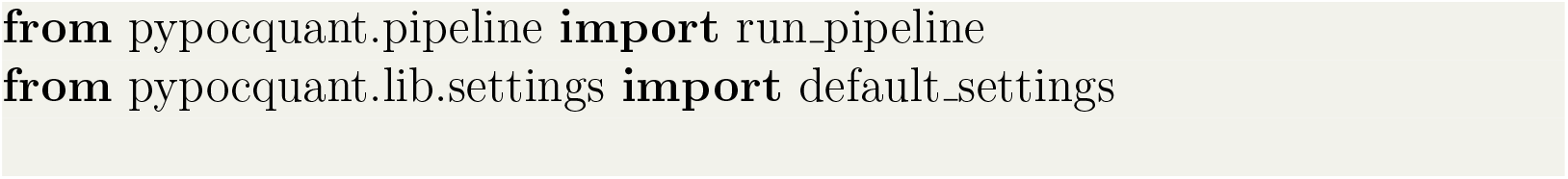
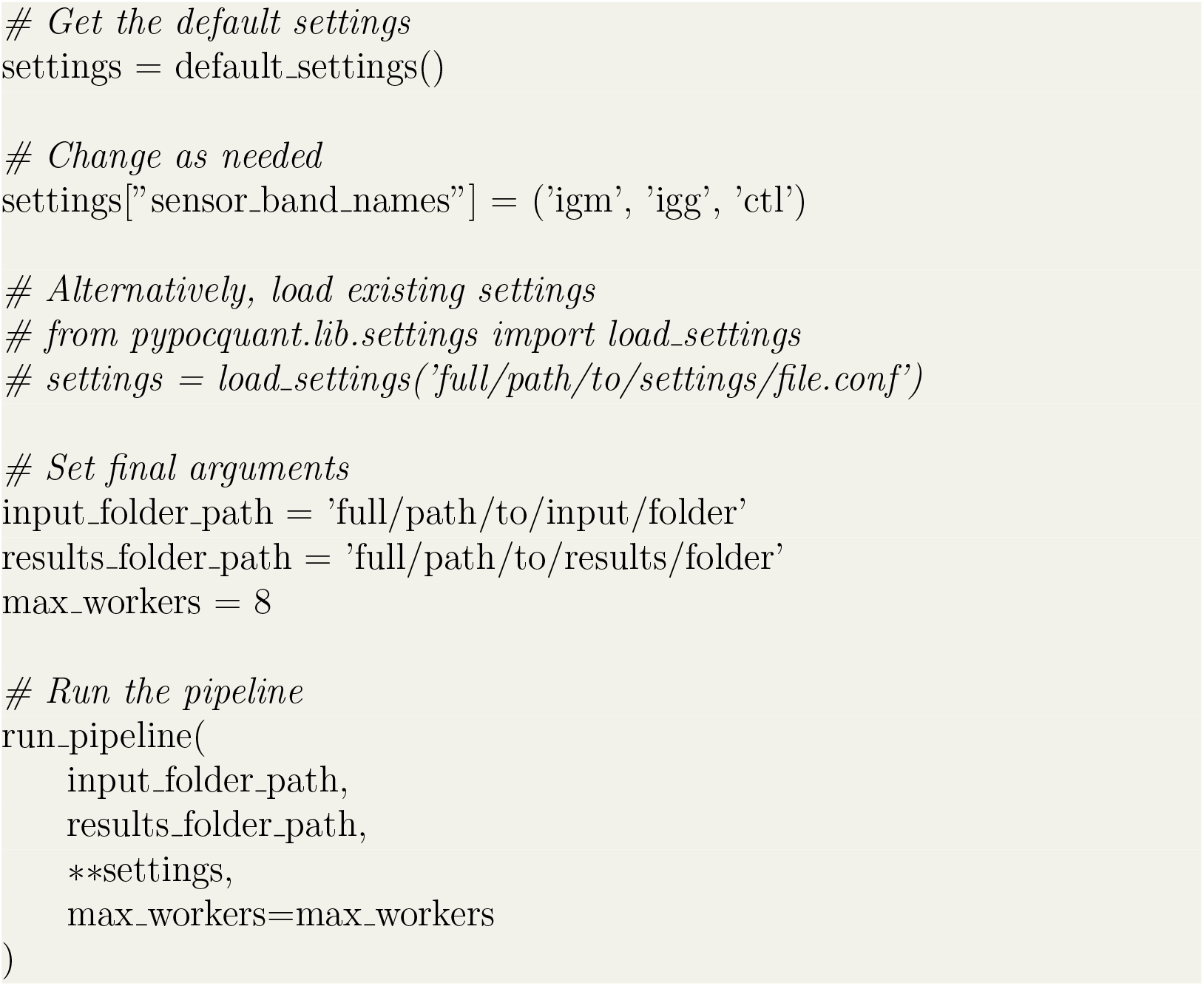

### 3.4. Example results

We will soon present a detailed, quantitative study of pyPOCQuant involving a large sample of images (manuscript in preparation). As an illustration of the performance of pyPOCQuant, we analyzed one image at random from each of 14 manufacturers in a set of 10,000 POCT images [22]. Figure 5 shows the extracted TL over the corresponding POCT sensor and area (as delimited by the four QR codes). We then compared the binary readout from pyPOCQuant with the visual detection of two researchers and found an accuracy of 1.00 (n=31 TLs on 14 POCTs). We summarise the results in the table at the bottom right panel of Figure 5. The Jupyter notebook used for the analysis can be found in the `examples` folder of the code repository. This first result shows that pyPOCQuant can identify TL from various POCT housing designs with high accuracy.

**Figure 5:**
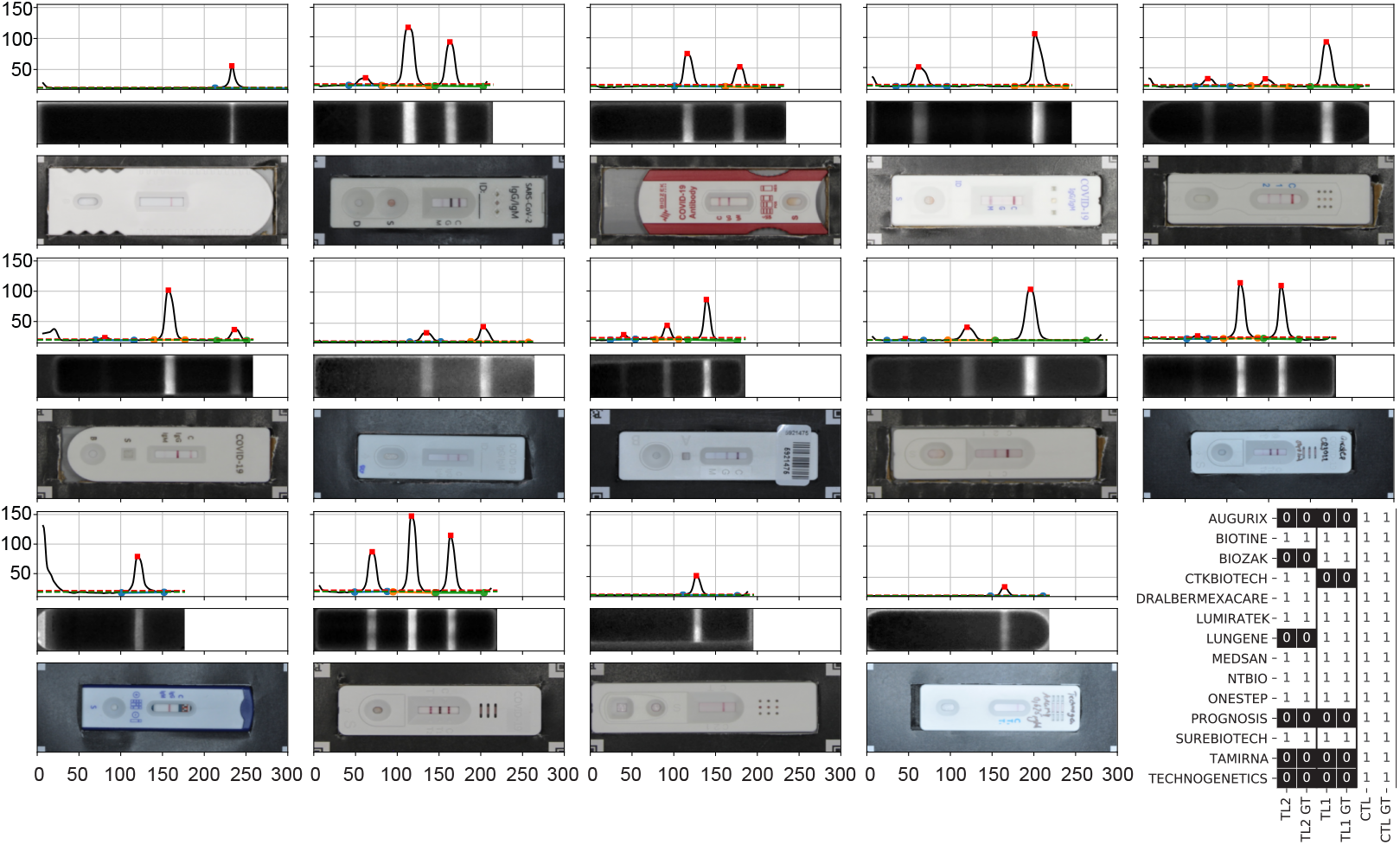
pyPOCQuant results for different POCT manufacturers. Top row depicts the background corrected signal and a red square marks the identified TL. Middle row depicts the detected sensor. Bottom row depicts the POCT inside the raw QR code box. The third entry in the first row shows an example of a POCT that was wrongly positioned in the QR code box and that is ultimately rotated for sensor extraction. The table shows the pyPOCQuant results compared to a manual ground truth for each TL.

## 4. Impact

We developed pyPOCQuant to analyse POCT images as a rapid and reliable alternative to visual scoring of these devices. While humans can quickly and easily spot the presence or absence of TLs of POCTs, this assessment is always subjective and poorly reproducible. Different operators may easily interpret weak test line signals differently and even the same operator may score the same weak test line as positive or negative under different conditions. Therefore, human assessment is neither reliable nor quantitative.

We propose pyPOCQuant as a valuable tool that generates quantitative and reproducible POCT results from lab evaluations all the way to large-scale testing campaigns. pyPOCQuant is not dependent on specialized equipment and can be used with POCTs from many different manufacturers. It therefore allows for straightforward quantification of cheap, mass-producible and readily available POCT using a consumer (or even a smartphone) camera and a mid-end computer.

The efficient scoring of POCTs by pyPOCQuant allows for simple validation, and improves their reliability in every day use. Scaling up is feasible as it can take advantage of multi-core computers to process images in parallel making it suited for large studies with thousands of associated images. As such, pyPOCQuant can help when large-scale testing using classic approaches such as PCR or ELISA may be hampered by limited access to reagents and laboratory equipment, such as during the global SARS-CoV-2 pandemic.

## 5. Conclusions

pyPOCQuant delivers accurate detection and precise quantification of test and control lines on any LFA. pyPOCQuant is available a library, commandline tool and graphical user-interface to fit into various analysis workflows. We developed it to reproducibly batch-process thousands of images of POCTs in parallel using multi-core computers or even clusters. We made pyPOC-Quant user-friendly by adding a graphical user interface to facilitate loading images, defining and testing analysis parameters on single images, and launching entire analysis campaigns with a few clicks. In addition to its execution from the CLI or GUI, pyPOCQuant can also run in Jupyter notebooks. Notebooks integrate analysis and documentation into elegant reports.

Image acquisition requirements are undemanding: consumer cameras provide acceptable images for analysis (6-12 MPixel cameras offer a good compromise between image resolution and size), and the provided imaging template makes pyPOCQuant compatible with a wide variety of POCT housing designs and sizes.

pyPOCQuant helps validating POCTs quickly and can be used to establish rapid field testing. It has already been used in several pilot SARS-CoV-2 related projects involving several thousand images. Since POCTs are not restricted to SARS-CoV-2 antigens, pyPOCQuant can be broadly used for large-scale diagnostics and studies of various other diseases.

## Data Availability

Data is freely available in the gitlab link

## 6. Conflict of Interest

The authors declare no competing interests.

### Contribution

**Andreas P. Cuny** Conceptualization, project administration, methodology, software, resources, investigation, data curation, formal analysis, validation, visualization, original draft preparation, writing, review and editing. **Aaron Ponti** Conceptualization, methodology, software, resources, investigation, data curation, validation, original draft preparation writing, review and editing. **Fabian Rudolf** Conceptualization, review and editing.

## Acknowledgements

We would like to thank Marvin Wyss and Talea Marty from the Fachhochschule Nordwestschweiz for valuable user feedback that helped us greatly improve the software and the user experience. We would like to thank Marc Obrist for providing us with a ring light for the photo box during the time of lockdown when resources were generally hard to get. FR is funded by the NCCR ‘Molecular Systems Engineering’.

