## Supplementary Material for "pyPOCQuant - A tool to automatically quantify Point-Of-Care Tests from images"

---

### *POCT template and mount*

To maximize reproducibility in comparing hundreds or thousands of POCT images, we designed a simple mount for the POCT that uses a printable template in A4 format. The template includes QR codes to assist the localization of the POCT (Suppl. Fig. S1). The top QR code (TL_P) allows to identify a POCT packaging with LOT number. The QR box with the QR codes TL, TR, BL, BR identifies the POCT on the image. The red grid helps to cut out an appropriate window for the POCT to be tested. The QR code R_G and L_G surround grey scale bars. The printed template is glued on a thick carton (approximately 5mm), and the central part is cut to create a pocket that keeps the POCTs stably in position (Suppl. Fig. S2).

### *Image acquisition setup*

Another critical aspect of the acquisition of images for robust analysis is homogeneous lighting conditions with no shadows that can bias the quantification and a fixed acquisition perspective. The most straightforward setup consists of the POCT mount described in the previous section immobilized on a table and an SLR camera mounted on a tripod. The image is shot in a dark room or a box. A ring light ensures constant, even light illumination; however, a normal flash works as well. For our studies, we built a photo box (Suppl. Fig. S3) from OpenBeam aluminum profiles, a black carton, a ball head mount for the SLR camera (Nikon D90 with a 24mm f2.8 Objective), a Doerr LED Macro Ring Light with 16 LEDs (Doerr LED-16 Ringlight, Art.-Nr. 371023) modified to be wall powered, and a few screws. Additionally, we built a stand from OpenBeam profiles, that allows a smartphone to be placed at a constant height above our template.

*Recommendation for improved POCT housing design*

POCTs feature high sensitivity and are cost effective, but their format does not lend itself for large-scale, batched analysis. A few dedicated readers exist, but they only work with specific manufacturers and models. pyPOCQuant relies on a specially-designed template and a configuration file to significantly extend the number of POCT types that can be analysed at large scale. Still, the process could be simplified even further if the POCT housing were modified to store specific metadata information in machine-readable form. The modifications would be minor. We suggest printing a QR code box around the LFA sensor for robust extraction of the area to quantify and add LOT, date, and possibly additional information as a data matrix directly on the housing. Any camera could read this information, thus making many currently essential computer vision algorithms in pyPOCQuant no longer necessary.

*Possible future developments*

While the typical use case for pyPOCQuant is during controlled clinical trials of pandemic (such as SARS-CoV-2), it could be deployed in the cloud for use by organizations or state authorities. The operator would interact with the cloud using a web application. The server would produce a unique, random, and anonymous ID that the operator would place on the imaging template during image acquisition. The acquired image would be uploaded and analyzed in the cloud. Results would then be added to a secured database and used to generate live statistics, plots, and reports that could inform the public and drive further actions.

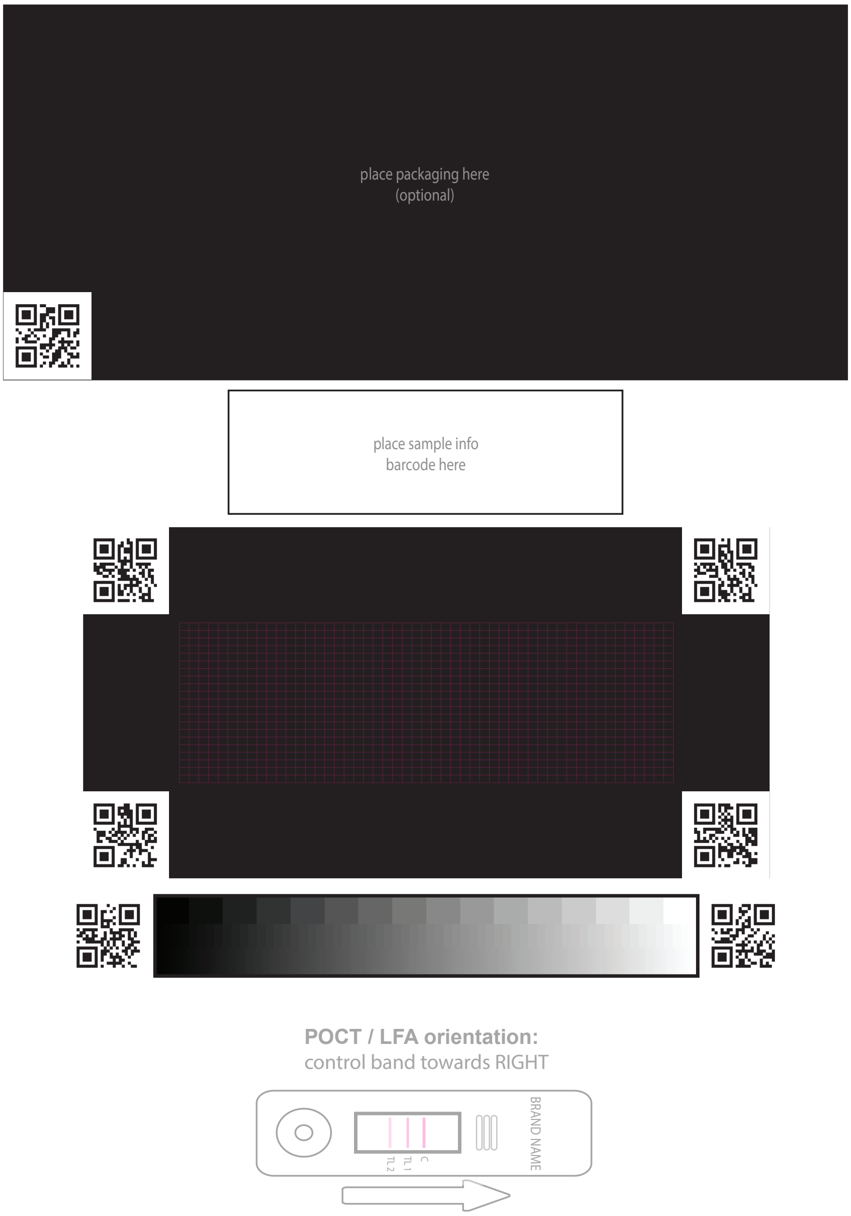


Figure S1: pyPOCQuant POCT recognition template.

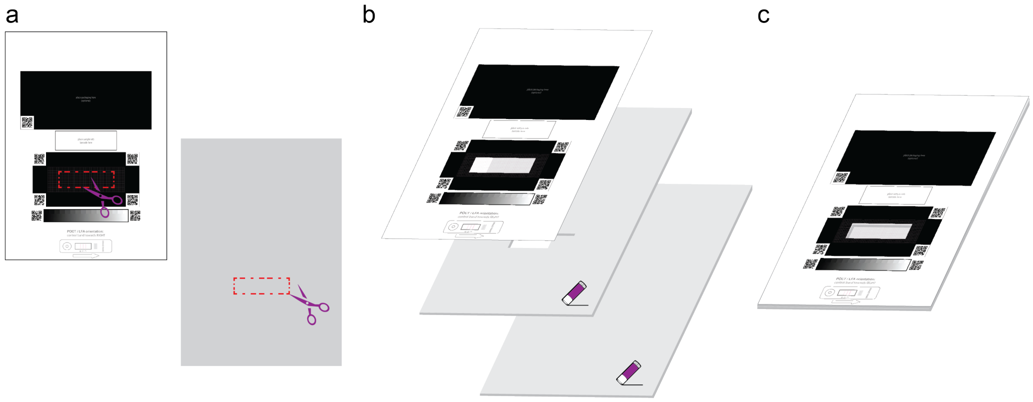


Figure S2: Assembly guide for a simple POCT mount. a) Print the POCT recognition template (Suppl. Fig. S1) in A4 format on a paper and cut out the area for the POCT and do to the same on a carton. c) Glue the template paper on the prepared carton and add a uncut carton below. c) Final mount

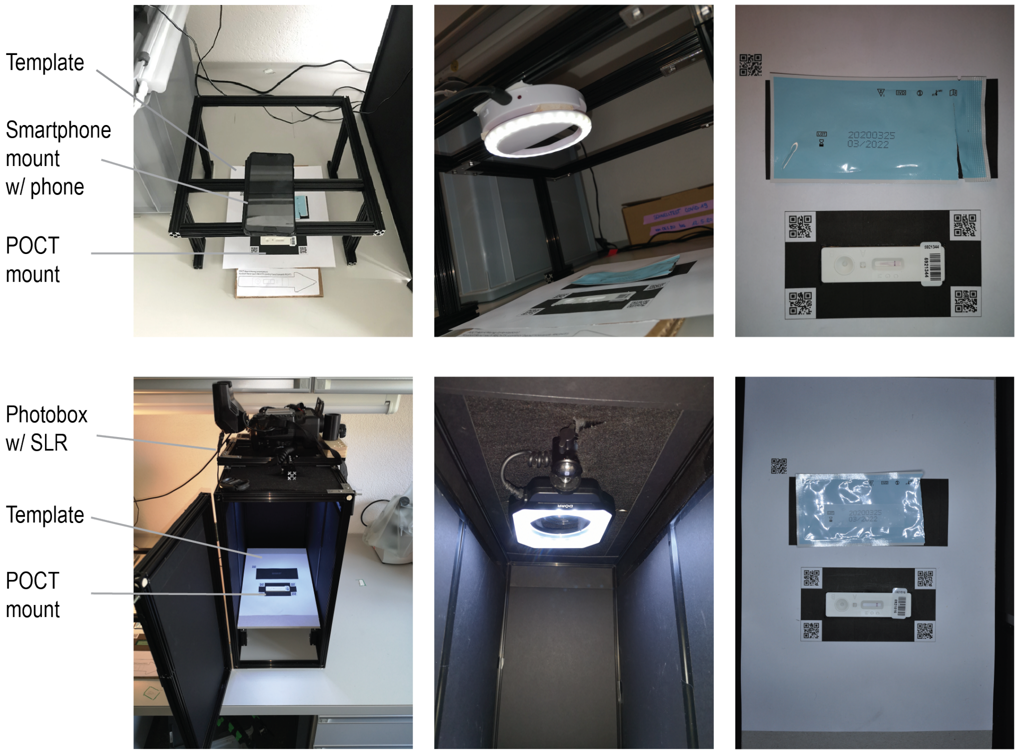


Figure S3: Depiction of our image acquisition setup. Top row, OpenBeam smartphone mount above initial version of the POCT template and mount. Bottom row, custom, photo box with controlled light conditions and fixed mounted camera.

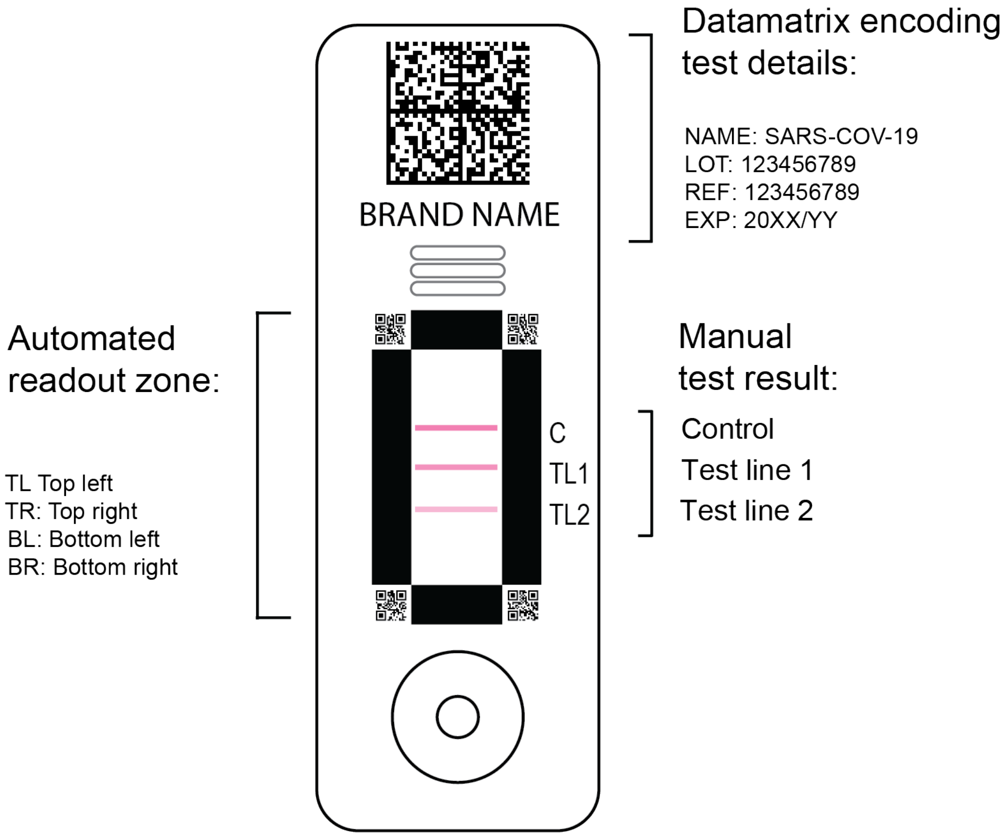


Figure S4: Suggestion for new machine readable POCT housing design.
